# A Targeted Geospatial Approach to COVID-19 Vaccine Delivery: Findings from the Johns Hopkins Hospital Emergency Department

**DOI:** 10.1101/2021.05.04.21255575

**Authors:** Sunil S. Solomon, Yu-Hsiang Hsieh, Richard E. Rothman, Oliver Laeyendecker, Mark Anderson, Shruti H. Mehta, Gavin Cloherty, Thomas C. Quinn

## Abstract

While COVID-19 vaccines have been shown to significantly decrease morbidity and mortality, there is still much debate about optimal strategies of vaccine rollout. We tested identity-unlinked stored remnant blood specimens of patients at least 18 years presenting to the Johns Hopkins Hospital emergency department (ED) between May to November 2020 for IgG to SARS-CoV-2. Data on SARS-CoV-2 RT PCR were available for patients who were tested due to suspected infection. SARS-CoV-2 infections was defined as either a positive IgG and/or RT-PCR. SARS-CoV-2 infection clustering by zipcode was analyzed by spatial analysis using the Bernoulli model (SaTScan software, Version 9.7). Median age of the 7,461 unique patients visiting the ED was 47 years and 50.8% were female; overall, 740 (9.9%) unique patients had evidence of SARS-CoV-2 infection. Prevalence of infection in ED patients by ZIP code ranged from 4.1% to 22.3%. The observed number of cases in ZIP code C was nearly double the expected (observed/expected ratio = 1.99; 95% CI: 1.62, 2.42). These data suggest a targeted geospatial approach to COVID vaccination should be considered to maximize vaccine rollout efficiency and include high-risk populations that may otherwise be subjected to delays, or missed.

## Introduction

COVID-19 vaccines decrease disease, mortality^1^ and infection,^2^ yet remain in limited supply. Even with expanded access, debate continues about how to optimize equitable distribution, particularly poignant in the face of a time-sensitive window of opportunity to curtail the pandemic amid threats from emerging variants.^3^ Persons visiting an emergency department (ED) during the pandemic are ill enough to require medical care and may represent communities at increased risk of COVID-19 associated morbidity and mortality,^4^ and who may benefit most from COVID vaccines.

## Methods

We tested identity-unlinked stored remnant blood specimens of patients ≥18 years presenting to the Johns Hopkins Hospital ED between May–November 2020 for IgG to SARS-CoV-2 using the Abbott Architect SARS-CoV-2 IgG Assay (Supplementary Methods). Data on SARS-CoV-2 RT PCR were available for patients who were tested due to suspected infection. Socio-demographic data namely age, sex at birth, race, ethnicity, residential zip code, month of visit, primary care doctor, and insurance payor were available. The protocol was approved by the Johns Hopkins Medicine Institutional Review Board (IRB00083646).

Evidence of SARS-CoV-2 positivity was defined as either having a positive RT-PCR and/or seropositivity to SARS-CoV-2 IgG of the stored specimen collected during the unique ED. Heat map of SARS-CoV-2 infection (IgG and/or RT-PCR) prevalence by ZIP code for the JHH ED catchment area in Baltimore City was created using Microsoft Excel Map function. Only zipcodes that contained at least 1% unique ED patients (n≥74) in the sample were plotted. SARS-CoV-2 infection clustering by zipcode was analyzed by spatial analysis using the Bernoulli model (SaTScan software, Version 9.7).

## Results

There were 24,615 ED visits by 17, 536 unique patients during the study period, of which remnant samples were available for 7,461 unique patients. Median age was 47 years and 50.8% were female (Table 1). 549 (7.4%) were positive for SARS-CoV-2 IgG and 277 (3.7%) clients tested RT-PCR positive during their visit; overall, 740 (9.9%) unique patients had evidence of SARS-CoV-2 infection. Infection was more common in Hispanic patients (36.1% vs. 4.2% in non-Hispanic Whites) and uninsured patients (18.5% vs. 9.0%).

**Table 1.** Socio-demographic characteristics of patients visiting the Johns Hopkins Hospital Emergency Department (May – November 2020) by Evidence of SARS-CoV-2 Infection

|  | Overall (n=7461) | Evidence of SARS-CoV-2 Infection (n= 740) | No Evidence of SARS-CoV-2 Infection (n= 6721) |
| --- | --- | --- | --- |
| Age in years* (IQR) | 47 (32, 61) | 45 (32, 59) | 47 (32, 61) |
| Biological sex at birth, n(%) |  |  |  |
| *Male* | 3668 (49.2) | 369 (49.9) | 3299 (49.1) |
| *Female* | 3791 (50.8) | 371 (50.1) | 3420 (50.9) |
| *Unknown* | 2 (0) | 0 | 2 (0) |
| Race/Ethnicity*, n(%) |  |  |  |
| *White, Non-Hispanic* | 2181 (29.2) | 92 (12.4) | 2089 (31.1) |
| *Black, Non-Hispanic* | 4245 (56.9) | 410 (55.4) | 3835 (57.1) |
| *Other race, Non-Hispanic* | 484 (6.5) | 39 (5.3) | 445 (6.6) |
| *Hispanic, any race* | 551 (7.4) | 199 (26.9) | 352 (5.2) |
| Zipcode of residence*, n(%) |  |  |  |
| *A* | 674 (9.0) | 70 (9.5) | 604 (9.0) |
| *B* | 450 (6.0) | 57 (7.7) | 393 (5.9) |
| *C* | 444 (6.0) | 99 (13.4) | 345 (5.1) |
| *D* | 395 (5.3) | 35 (4.7) | 360 (5.4) |
| *E* | 351 (4.7) | 40 (5.4) | 311 (4.6) |
| *F* | 322 (4.3) | 37 (5.0) | 285 (4.2) |
| *G* | 296 (4.0) | 28 (3.8) | 268 (4.0) |
| *H* | 207 (2.8) | 17 (2.3) | 190 (2.8) |
| *I* | 204 (2.7) | 23 (3.1) | 181 (2.7) |
| *J* | 140 (1.9) | 21 (2.8) | 119 (1.8) |
| *Other†* | 3978 (53.3) | 313 (42.3) | 3665 (54.5) |
| ZCTA Poverty Level*‡, n(%) |  |  |  |
| <10% | 1944 (26.1) | 148 (20.0) | 1796 (26.7) |
| 10% to <20% | 2329 (31.2) | 262 (35.4) | 2067 (30.7) |
| 20% to <30% | 1859 (24.9) | 192 (25.9) | 1667 (24.8) |
| ≥30% | 1164 (15.6) | 124 (16.8) | 1040 (15.5) |
| Unavailable | 165 (2.2) | 14 (1.9) | 151 (2.3) |
| Primary Care Physician*, n(%) |  |  |  |
| *Yes* | 4694 (62.9) | 426 (57.6) | 4268 (63.5) |
| *No* | 2767 (37.1) | 314 (42.4) | 2453 (36.5) |
| Self-Pay or No Primary Payor*, n(%) |  |  |  |
| *Yes* | 733 (9.8) | 136 (18.4) | 597 (8.9) |
| *No* | 6728 (90.2) | 604 (81.6) | 6124 (91.1) |

\* $p < 0.05$
† other includes 649 zipcodes and none/missing zipcodes (n=131)
‡ ZIP code tabulation area (ZCTA) data from the 2018 American Community Survey of 5-year estimate for proportion living below the poverty level

Prevalence of infection (IgG and/or RT-PCR) in ED patients by ZIP code ranged from 4.1% to 22.3% (Figure 1). The observed number of cases in Zipcode C was nearly double the expected (observed/expected ratio = 1.99; 95% CI: 1.62, 2.42).

**Figure 1.**
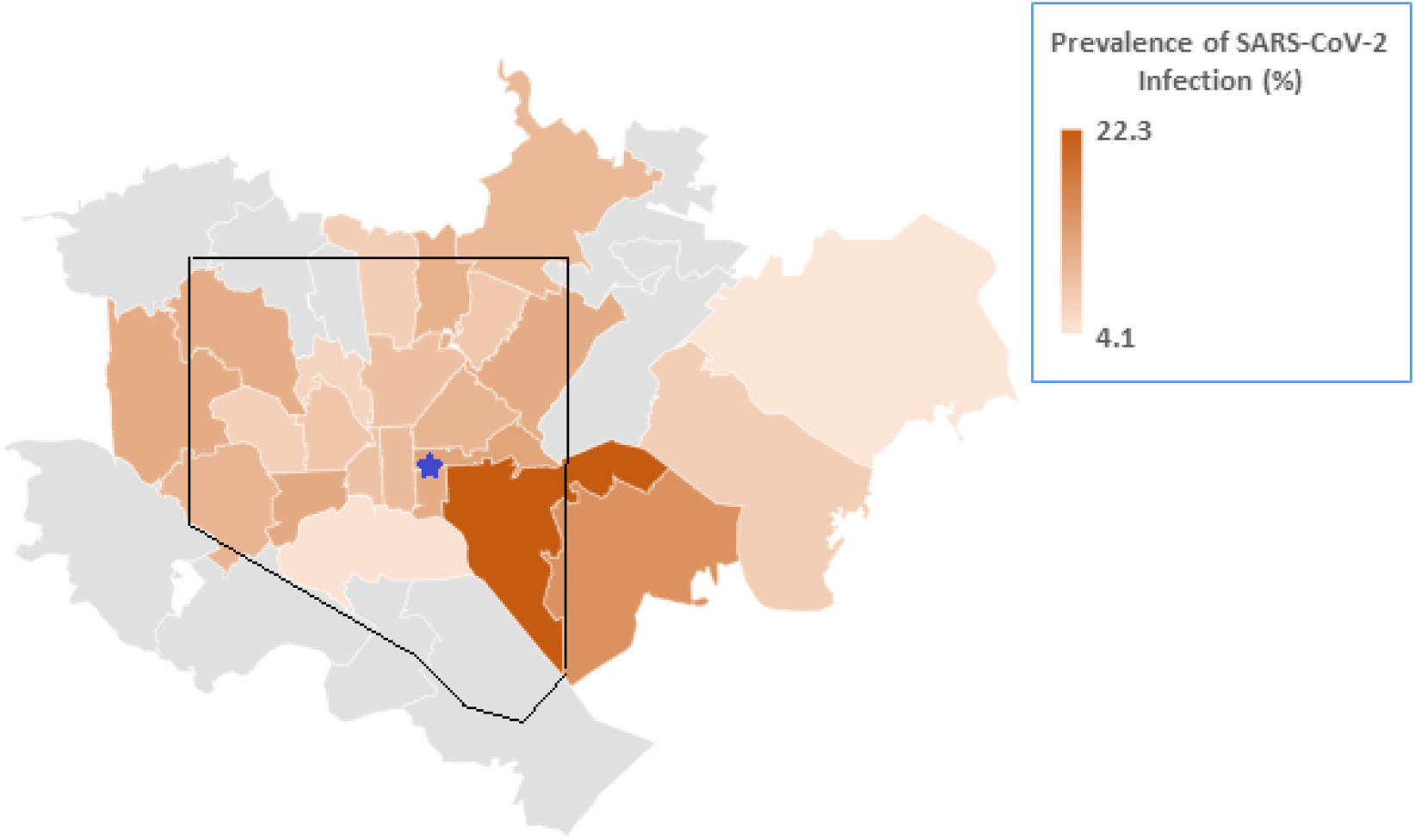
SARS-CoV-2 prevalence by zipcode among patients visiting the Johns Hopkins Hospital Emergency Department between May through November 2021. Patients were classified as having SARS-CoV-2 if they either had a positive SARS-CoV-2 RT-PCR during their ED visit and/or had a stored specimen available that tested positive for the presence of SARS-CoV-2 IgG. Blue star indicates the location of study ED. The black line indicates the Baltimore City and Baltimore County boundary line.

## Discussion

These data show that in addition to demographic and clinical criteria (e.g., age, comorbidities), residential ZIP code is predictive of SARS-CoV-2 infection and should be used to prioritize vaccine distribution without stigmatizing or targeting racial, ethnic, or other groups. Focusing on ZIP codes, which in this population are known to be in high density, overcrowded, poorer locations with multi-generational homes, could interrupt transmission. Supply and distribution chains could be streamlined given known population denominators for ZIP codes. The approval of a single-dose vaccine requiring only refrigeration lends itself to a field-based delivery system. Further, immunizing full households (vs. subsets) accounts for SARS-CoV-2 transmission dynamics^5^ and allows for easier adoption of CDC guidelines for vaccinated individuals.^6^ Finally, ZIP codes reflect socially connected populations (i.e. grass root community-based organizations), who can champion vaccines and reduce hesitancy.

If SARS-CoV-2 infection status of ED patients is unavailable, and to account for limited catchment area of EDs and multiple EDs in a city, residential ZIP code of people who have either died of COVID-19 or been hospitalized could be used to formulate a similar targeted approach. Even with heterogeneous specimen availability or locally collected hospitalization data, a targeted geospatial approach to COVID vaccination should be considered to maximize vaccine rollout efficiency and include high-risk populations that may otherwise be subjected to delays, or missed.

## Data Availability

The data that support the findings of this study are available from the corresponding author upon reasonable request.

## Acknowledgements

This study was partly funded by Abbott Laboratories, USA.

## REFERENCES

1. Kim JH, Marks F, Clemens JD. Looking beyond COVID-19 vaccine phase 3 trials. Nat Med 2021;27(2):205–211.

2. Amit S, Regev-Yochay G, Afek A, Kreiss Y, Lesham E. Early rate reductions of SARS-CoV-2 infection and COVID-19 in BNT162b2 vaccine recipients. Lancet 2021;397(10277):875–77.

3. Altmann DM, Boyton RJ, Beale R. Immunity to SARS-CoV-2 variants of concern. Science 2021;371(6534):1103–1104.

4. Boserup B, McKenney M, Elkbuli A. The impact of the COVID-19 pandemic on emergency department visits and patient safety in the United States. Am J Emerg Med 2020;38(9):1732–1736.

5. Madewell ZJ, Yang Y, Longini, Jr IM, Halloran E, Dean NE. Household Transmission of SARS-CoV-2: A Systematic Review and Meta-analysis. JAMA Network Open 2020;3(12): e2031756.

6. Interim Public Health Recommendations for Fully Vaccinated People. Atlanta, GA: Centers for Disease Control and Prevention (CDC), March 2021 (https://www.cdc.gov/coronavirus/2019-ncov/vaccines/fully-vaccinated-guidance.html)

