## Supplementary File 1 for "A Targeted Geospatial Approach to COVID-19 Vaccine Delivery: Findings from the Johns Hopkins Hospital Emergency Department"

**Methods**

*Study Setting and Population*

An identity-unlinked seroprevalence study was conducted at Johns Hopkins Hospital Emergency Department (JHH ED) during May 1, 2020 – July 4, 2020 and August 30, 2020 – November 9, 2020. All available remnant blood from hematology samples from ED patients were collected during the study period. For each sample, a unique study code was assigned, processed, and stored at −80°C. For all samples, basic patient sociodemographic characteristics (age, sex, race, ethnicity, residential zip code, month of visit, primary care doctor, and insurance payor) and SARS-CoV-2 PCR testing and the result, were abstracted from medical records, and all identifiers and protected health information removed from the dataset.

The study population comprised a subset of patients who visited the Johns Hopkins Hospital Emergency Department during the study period. During this time, there were 24,615 visits to the Emergency Department by 17,536 unique patients. Identity-unlinked remnant specimens were collected from 9,434 patients of which 9,227 were sent for testing. Specimens were not sent for testing if they had insufficient volume for testing. Of the 9,227 sent for testing, samples from 7,461 unique patients were found to have sufficient volume for SARS-CoV-2 IgG testing and were included in this sample after excluding specimens. Multiple samples from the same visit, patients ≤18 years of age and samples from non-emergency department visits were excluded.

*Laboratory procedures*

All stored specimens were tested for the presence of SARS-CoV-2 IgG using the FDA EUA approved Abbott ARCHITECT SARS-CoV-2 IgG (List 06R86) assay as per the ARCHITECT operations manual and assay package insert instructions. Briefly, the SARS-CoV-2 IgG assay is an automated Chemiluminescent Microparticle Immunoassay (CMIA) used for the qualitative detection of IgG antibodies directed against the SARS-CoV-2 Nucleocapsid protein. Assay results are measured in Relative Light Units (RLU) and reported as an index value of the ratio of specimen to calibrator RLU signal (S/C). Index values ≥1.4 S/C indicate a SARS-CoV-2 IgG seropositive result and were used to determine evidence of prior SARS-CoV-2 infection. Patients in the emergency department with suspected SARS-CoV-2 infection were tested for active SARS-CoV-2 infection using RT-PCR under ED physician’s clinical discretion.

*Statistical Analysis*

Evidence of SARS-CoV-2 infection was defined as any SARS-CoV-2 PCR positive result during the study period or seropositivity of SARS-CoV-2 IgG of the specimen collected during the unique ED visit or the last ED visit for those patients with multiple ED visits and specimens. Race/ethnicity of subjects were categorized into Hispanic/Latino, Non-Hispanic White, Non-Hispanic Black, or Non-Hispanic Other Race. Residential zip codes of subjects were categorized by the most common individual 10 zip codes and the remaining zip codes categorized as ‘other’. Residential zip code was also linked to ZIP code tabulation area (ZCTA) data from the 2018 American Community Survey of 5-year estimates for proportion living below the poverty level which was categorized into <10%, 10-20%, 20-30%, ≥ 30%, or missing.^1,2^ All data analyses were performed using SAS V.9.4 (SAS Institute Inc., Cary, North Carolina) and a two-sided p-value less than 0.05 was considered statistically significant. Heat map of SARS-CoV-2 infection (IgG and/or RT-PCR) prevalence by zipcode for the JHH ED catchment area in Baltimore City was created using Microsoft Excel Map function. Only zipcodes that contained at least 1% unique ED patients (n≥74) in the sample were plotted. SARS-CoV-2 infection clustering by zipcode was analyzed by spatial analysis using the Bernoulli model (SaTScan software, Version 9.7).^3^

**REFERENCES:**

1. United States Census Bureau. Zip code tabulation areas (ZCTA). Available at: https://www.census.gov/programs-surveys/geography.html. Accessed February 9, 2021.
2. United States Census Bureau. US Census Bureau American Community Survey. 2018 American Community Survey 5 year estimates, tables B03002, S1701, and B01003. Available at: https://data.census.gov/cedsci/. Accessed February 9, 2021
3. Software: Kulldorff M. and Information Management Services, Inc. SaTScanTM v9.7: Software for the spatial and space-time scan statistics. http://www.satscan.org/, 2021.
